# Comparison of Facial Feature Grading by an AI-Based System Versus Human Expert Grading on Images Under Standardized Clinical and At-Home Settings

**DOI:** 10.64898/2026.09.30.26364271

**Authors:** Daniil Danko, Timur Tlyachev, Konstantin Kiselev, Lydia Bousseksou, Etienne Camel, Georgios Stamatas, Anastasia Georgievskaya

## Abstract

Digital photography remains the gold standard for dermatological assessment, but reliance on standardized stationary image-capture systems limits trial scalability, whereas smartphones combined with computer vision artificial intelligence (AI) could enable hybrid and decentralized clinical trials (DCTs). This study assessed the facial feature grading of an AI-based system against expert panel consensus across three settings with decreasing acquisition control: Standard Studio Imaging (SSI), Clinic Smartphone Imaging (CSI), and Home Smartphone Imaging (HSI). Expert inter-rater reliability was good to excellent (median ICC 0.84–0.90), and the system showed high operator-independent test-retest repeatability across independently captured images (median ICC 0.97–0.98). Agreement with experts was maintained across all settings for macro-geometric features (ptosis *ρ* = 0.74–0.81, nasolabial fold *ρ* = 0.68–0.79) and for discrete pigmentation and inflammatory lesions (pigmented spot density *ρ* = 0.62–0.76), declined for fine-texture features under uncontrolled acquisition (forehead wrinkles *ρ* = 0.76 to 0.48), and remained limited for color-dependent features irrespective of setting. These findings support the AI-based system as an objective grading tool for a defined subset of facial endpoints in hybrid and DCTs in dermatology.

## Introduction

Skin imaging techniques have a long history in dermatological and cosmetological sciences, dating back to 1848 when the first illustrative photography was published by Dr. Samuel P. Hullihen (Neuse et al. 1996). Since then, a plethora of imaging techniques ranging from photography to multispectral microscopy has been developed and implemented by scientists (Chekanov et al. 2024; Gaurav et al. 2024). Nevertheless, digital photography remains one of the most convenient methods of scientific communication among dermatologists. This approach is crucial for accurate diagnostics, tracking aging progression, and treatment efficacy. As a “gold standard” for dermatological assessment, grading with the help of various scales, dermatological atlases, and scoring systems by a certified expert have been used (Bazin and Flament 2010; Bhor and Pande 2006). However, this type of evaluation can be subject to bias due to the observer effect, related to the assessor, and measurement bias, resulting from instrument-dependent variability and image acquisition standardization (e.g., illumination conditions, setting inhomogeneity or built-in image post-processing, etc.) (Dehon and Brédart 2001; George and Hole 1995; Georgievskaya et al. 2025; Ilic et al. 2022).

To mitigate such bias in medical imaging, highly standardized objective systems, such as VISIA^®^(Canfield Imaging Systems, Fairfield, NJ, USA) or ColorFace^®^ (QIMA Newtone, Lyon, France), have been developed and implemented in clinical practice and laboratory settings. The resulting images have highly standardized setups, such as head positioning, illumination angle and intensity, and camera settings including high resolution, combined with the possibility of image acquisition at different polarization or fluorescence conditions. However, this high-quality skin assessment can only be achieved within a clinical setting or in specially designed imaging booths. This limits the scalability of studies, narrowing the included cohort to local participants (Skea et al. 2019).

Nowadays, smartphones with high-resolution cameras are widespread, and combining them with the recent advances in artificial intelligence (AI) in dermatological and cosmetological sciences (Choy et al. 2023; Lee et al. 2025; Lei et al. 2025; Wilczok 2025) presents a unique opportunity for a decentralized mode of clinical trials. Recent technological research has enabled the development of a wide repertoire of AI-powered computer vision (CV) algorithms for the evaluation of skin features relating to age-dependent features (Georgievskaya et al. 2024b) such as hyperpigmented spots, wrinkles, acne, etc. (Tobar et al. 2024) or for the diagnosis of dermatological disease (Escalé-Besa et al. 2024). The possibility to use such methods to collect data remotely opens the opportunity to design studies at a larger scale increasing the number of participants. At the same time, it introduces variabilities in image acquisition due to uncontrolled conditions including different cameras, illumination sources, imaging geometries, head posture, and others (Georgievskaya et al. 2025). A rigorous validation process of such technologies against gold standards, such as expert assessment in controlled environments, is needed to ensure the accuracy and applicability of such AI-driven assessments for clinical-quality trials.

The aim of this study was to assess the performance of an AI-based system in terms of grading of facial features against grading by trained assessors in three different study settings: fully controlled illumination and head positioning and high-resolution images with a standardized system (SSI), smartphone camera-based images captured by a trained technician in a clinical setting (CSI), and smartphone images captured by participants in the highly variable environments of their homes (HSI). By analyzing the reliability and repeatability of the performance of the system across various environments, this study aims to establish the suitability of the AI-based system as an image capturing and grading tool that can be useful in decentralized clinical studies and remote facial analysis.

## Results

### Reliability and Reproducibility of the Human Experts Grading

To establish a baseline for ground truth, inter-rater reliability was evaluated between trained assessors. For most of the parameters, the intraclass correlation coefficient (ICC) indicated a good to excellent reliability of the grades provided by experts for all study settings (Table 1). Median reliability of the consensus for the SSI setting was 0.89 (IQR 0.83–0.91), for CSI 0.84 (IQR 0.81–0.90), and for HSI 0.90 (IQR 0.77–0.91). Overall, median reliability allows us to proceed with the following analysis and use averaged grades from 3 trained assessors for data comparison.

**Table 1.** Experts reliability within the experimental settings. Agreement scores among 3 expert assessors during the grading of 13 different facial features on photos for evaluation of an AI-powered skin analysis system in 3 settings (SSI – high-resolution images captured in a highly standardized environment, CSI – smartphone images captured by a trained technician in a standardized environment, HSI – smartphone images captured by participants in their homes). ICC(3, *k*) – two-way mixed model, average measures, consistency intraclass correlation coefficient; IQR – interquartile range.

| Facial feature | SSI | CSI | HSI |
| --- | --- | --- | --- |
| | ICC(3, $k$) | ICC(3, $k$) | ICC(3, $k$) |
| *Macro-geometric* |  |  |  |
| Ptosis, lower face | 0.92 | 0.95 | 0.90 |
| Nasolabial fold | 0.93 | 0.95 | 0.92 |
| Eye bags | 0.83 | 0.84 | 0.90 |
| Lacrimal groove | 0.88 | 0.81 | 0.91 |
| *Fine-texture* |  |  |  |
| Wrinkles, forehead | 0.96 | 0.93 | 0.93 |
| Wrinkles, under eye | 0.91 | 0.86 | 0.90 |
| Pores | 0.87 | 0.81 | 0.92 |
| *Discrete pigmentary / inflammatory* |  |  |  |
| Pigmented spots, density | 0.80 | 0.84 | 0.77 |
| Breakouts | 0.89 | 0.90 | 0.90 |
| *Color-dependent* |  |  |  |
| Diffused redness | 0.89 | 0.82 | 0.64 |
| Color uniformity | 0.67 | 0.62 | 0.40 |
| Dark circles, vascular | 0.69 | 0.64 | 0.72 |
| Dark circles, pigmentary | 0.90 | 0.86 | 0.91 |
| Median (IQR) | 0.89 (0.83–0.91) | 0.84 (0.81–0.90) | 0.90 (0.77–0.91) |

Macro-geometric features “Ptosis, lower face” and “Nasolabial fold”, together with the fine-texture feature “Wrinkles, forehead”, showed the highest agreement, with coefficients above 0.90 in all study settings. At the same time, color-dependent facial features “Diffused redness”, “Color uniformity”, and “Dark circles, vascular” showed higher variability in the grades, especially in the HSI setting (“Diffused redness” with 0.64 and “Color uniformity” with 0.40).

To assess the consistency of the experts’ grading between various study settings, correlation coefficients (Spearman’s *ρ* and confidence interval) and the intraclass correlation coefficient for absolute agreement, ICC(3, 1), were used (Table 2).

**Table 2.**
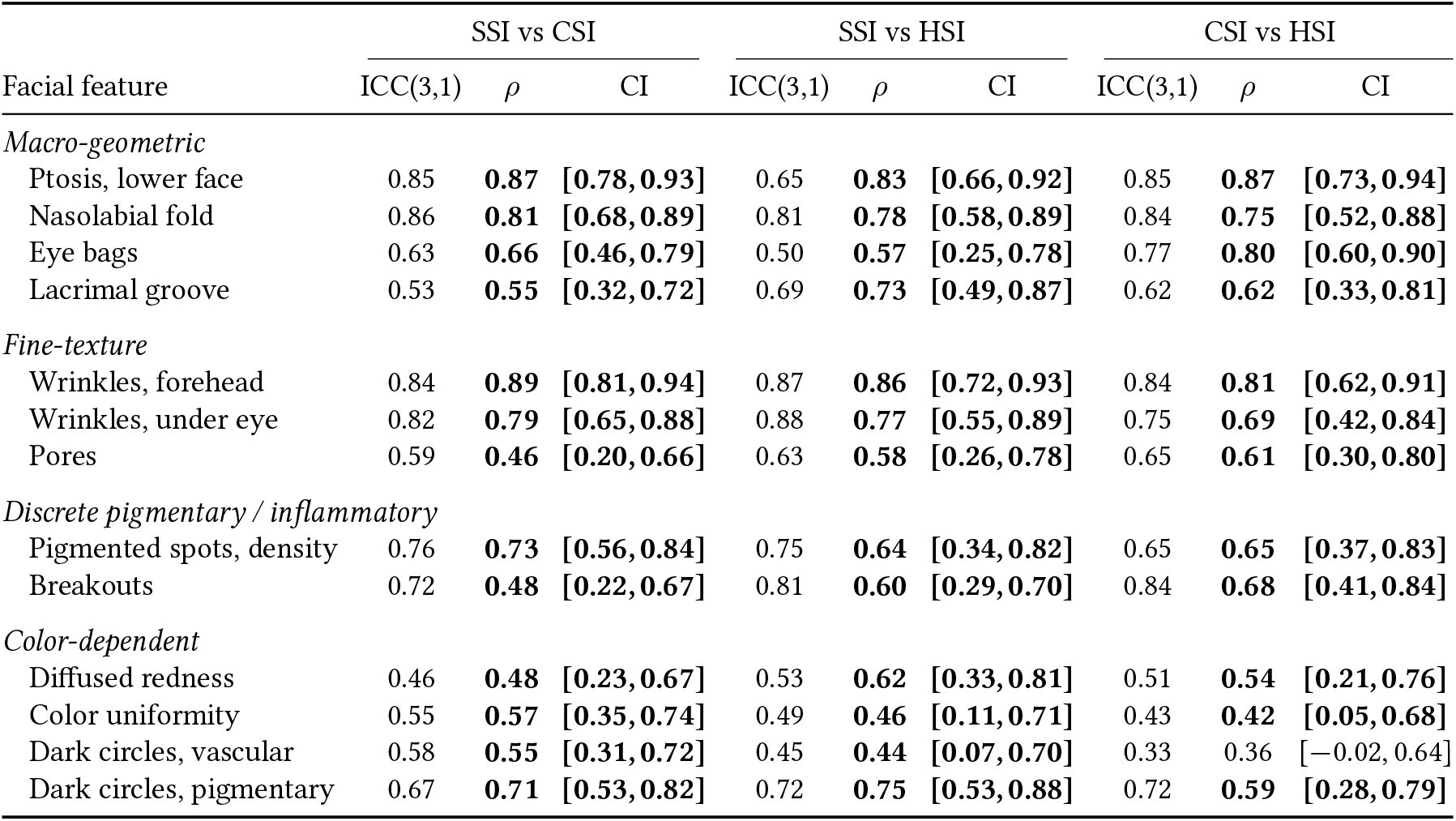
Reproducibility of the expert panel’s grades across three study settings (SSI – high-resolution images captured in a highly standardized environment, CSI – smartphone images captured in a standardized environment by a trained technician, HSI – smartphone images captured by participants in their homes) for 13 different facial features on photos. ICC – intraclass correlation coefficient; *ρ* – Spearman’s correlation coefficient; CI – 95% confidence interval. Values with non-zero confidence intervals are indicated in bold.

The experts’ grading showed high reproducibility of the results comparing SSI and smartphone photos (CSI and HSI study settings) for macro-geometric and fine-texture features: “Wrinkles, forehead” and “Nasolabial fold” with an ICC in the range 0.81–0.87. Comparing the two smartphone-based image capture settings (HSI vs CSI), reproducibility was high for the majority of the parameters. The “Ptosis” parameter showed high reproducibility across all settings (ICC *≥* 0.65) with high values for CSI vs HSI comparison (ICC = 0.85). “Breakouts” (ICC = 0.84), “Eye bags” (ICC = 0.77), and “Pigmented spots, density” (ICC = 0.65) showed moderate to strong agreement.

In contrast, agreement for color-dependent features was slightly lower. The “Dark circles, vascular” parameter presented poor reproducibility for the smartphone-based settings comparison (ICC = 0.33). For comparison, in the SSI study setting, the reproducibility was moderate (ICC 0.45–0.58). The “Diffused redness” parameter also showed moderate agreement in reproducibility of the results between study settings.

SSI, the high-resolution in-clinic setting, demonstrated better performance in grading color-dependent features. At the same time, macro-geometric feature grading was robust across all study settings for expert assessment, regardless of who captured the image (Table S1.1).

Taking into account analysis of the grades assigned by the expert assessors, we can conclude that the provided annotation can be a reliable ground truth for the assessment of the AI-powered system’s performance.

### Repeatability and Reproducibility of the AI-based System

To evaluate the repeatability of the analysis provided by the AI-based system, three consecutive images were taken and analyzed. Overall, the system demonstrated high repeatability (*>*0.9) in controlled and uncontrolled environments, as shown in Table 3. The results of the statistical analysis revealed that the median intraclass correlation coefficient for the CSI setting was 0.98 (IQR 0.96–0.98), and for the HSI setting, this coefficient was 0.97 (IQR 0.96–0.98).

**Table 3.** Repeatability of the AI-based facial skin analysis system across two study settings for various facial features. Agreement scores between three images of the AI analysis of different facial features on photos for evaluation of the AI-powered skin analysis system in two settings (CSI – smartphone images captured by a trained technician in a standardized environment, HSI – smartphone images taken by participants in their homes). ICC(3, *k*) – two-way mixed model, average measures, consistency intraclass correlation coefficient; IQR – interquartile range.

| Facial feature | CSI | HSI |
| --- | --- | --- |
| | ICC(3, $k$) | ICC(3, $k$) |
| *Macro-geometric* |  |  |
| Ptois, lower face | 0.98 | 0.99 |
| Nasolabial fold | 0.97 | 0.96 |
| Eye bags | 0.95 | 0.96 |
| Lacrimal groove | 0.95 | 0.95 |
| *Fine-texture* |  |  |
| Wrinkles, forehead | 0.98 | 0.99 |
| Wrinkles, under eye | 0.96 | 0.94 |
| Pores | 0.96 | 0.98 |
| *Discrete pigmentary / inflammatory* |  |  |
| Pigmented spots, density | 0.98 | 0.98 |
| Breakouts | 0.97 | 0.97 |
| *Color-dependent* |  |  |
| Diffused redness | 0.98 | 0.98 |
| Color uniformity | 0.98 | 0.99 |
| Dark circles, vascular | 0.98 | 0.95 |
| Median (IQR) | 0.98 (0.96–0.98) | 0.97 (0.96–0.98) |

The highest repeatability was detected for the aging-related skin features: “Wrinkles, forehead” (0.98 CSI, 0.99 HSI), “Ptosis” (0.98 and 0.99), and “Pigmented spots, density” (0.98 and 0.98). Notably, high repeatability was observed for the rating of “Color uniformity” by the AI system (ICC = 0.99 HSI).

To assess reproducibility of the AI system results between the 3 different study settings, varying in the resolution of the camera and imaging geometry and settings (including light conditions), agreement (ICC(3, 1) and correlation coefficients) has been calculated and the results are shown in Table 4.

**Table 4.**
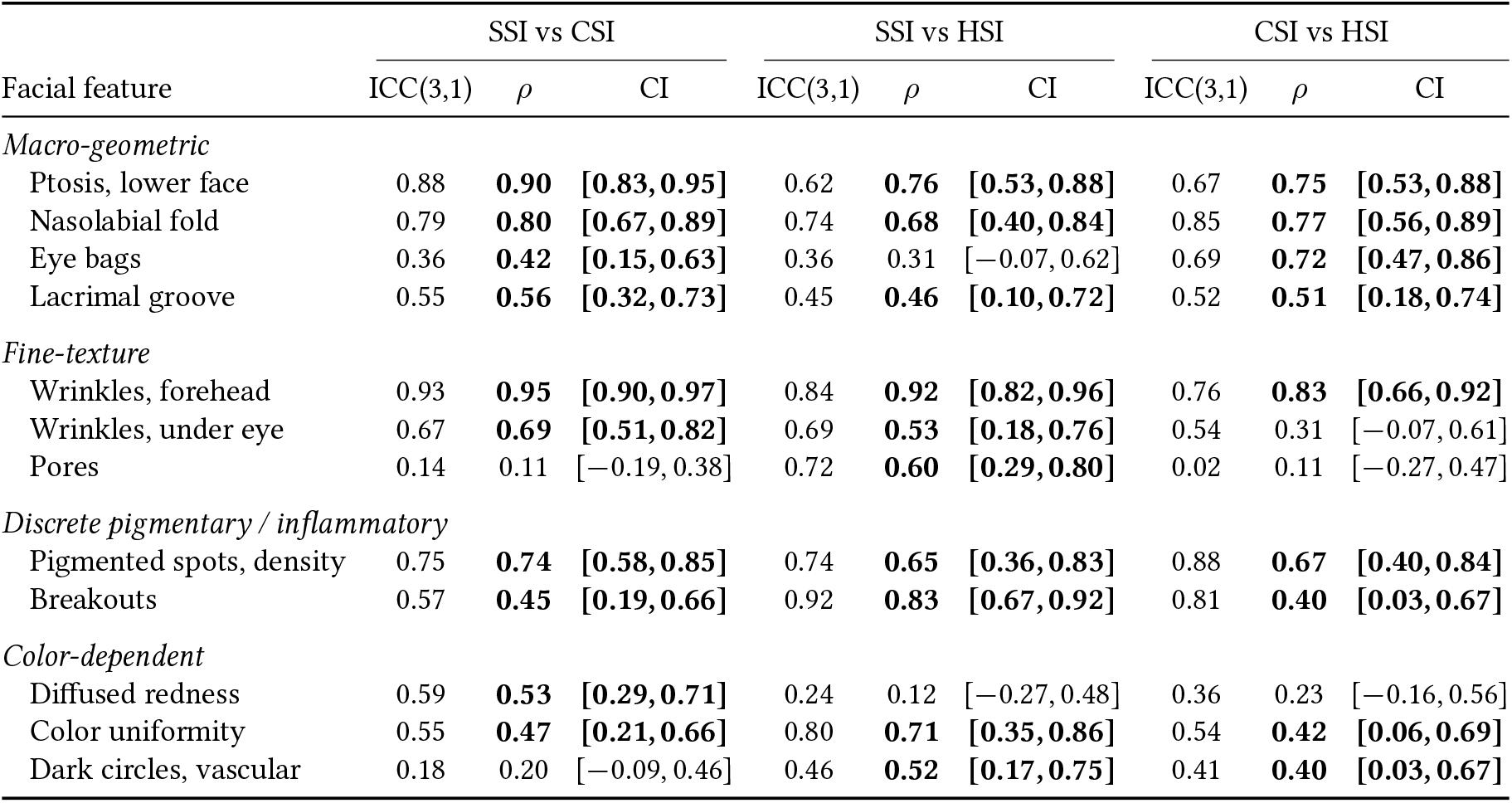
Reproducibility of AI-predicted facial skin analysis grades provided by the AI-based system across three study settings (SSI – high-resolution images captured in a highly standardized environment, CSI – smartphone images captured by a trained technician in a standardized environment, HSI – smartphone images captured by participants in their homes) for different facial features on photos. ICC – intraclass correlation coefficient; *ρ* – Spearman’s correlation coefficient; CI – 95% confidence interval. Values with non-zero confidence intervals are indicated in bold.

| Facial feature | SSI vs CSI |  |  | SSI vs HSI |  |  | CSI vs HSI |  |  |
| --- | --- | --- | --- | --- | --- | --- | --- | --- | --- |
| | ICC(3,1) | $\rho$ | CI | ICC(3,1) | $\rho$ | CI | ICC(3,1) | $\rho$ | CI |
| *Macro-geometric* |  |  |  |  |  |  |  |  |  |
| Ptosis, lower face | 0.88 | **0.90** | **[0.83, 0.95]** | 0.62 | **0.76** | **[0.53, 0.88]** | 0.67 | **0.75** | **[0.53, 0.88]** |
| Nasolabial fold | 0.79 | **0.80** | **[0.67, 0.89]** | 0.74 | **0.68** | **[0.40, 0.84]** | 0.85 | **0.77** | **[0.56, 0.89]** |
| Eye bags | 0.36 | **0.42** | **[0.15, 0.63]** | 0.36 | 0.31 | [-0.07, 0.62] | 0.69 | **0.72** | **[0.47, 0.86]** |
| Lacrimal groove | 0.55 | **0.56** | **[0.32, 0.73]** | 0.45 | **0.46** | **[0.10, 0.72]** | 0.52 | **0.51** | **[0.18, 0.74]** |
| *Fine-texture* |  |  |  |  |  |  |  |  |  |
| Wrinkles, forehead | 0.93 | **0.95** | **[0.90, 0.97]** | 0.84 | **0.92** | **[0.82, 0.96]** | 0.76 | **0.83** | **[0.66, 0.92]** |
| Wrinkles, under eye | 0.67 | **0.69** | **[0.51, 0.82]** | 0.69 | **0.53** | **[0.18, 0.76]** | 0.54 | 0.31 | [-0.07, 0.61] |
| Pores | 0.14 | 0.11 | [-0.19, 0.38] | 0.72 | **0.60** | **[0.29, 0.80]** | 0.02 | 0.11 | [-0.27, 0.47] |
| *Discrete pigmentary / inflammatory* |  |  |  |  |  |  |  |  |  |
| Pigmented spots, density | 0.75 | **0.74** | **[0.58, 0.85]** | 0.74 | **0.65** | **[0.36, 0.83]** | 0.88 | **0.67** | **[0.40, 0.84]** |
| Breakouts | 0.57 | **0.45** | **[0.19, 0.66]** | 0.92 | **0.83** | **[0.67, 0.92]** | 0.81 | **0.40** | **[0.03, 0.67]** |
| *Color-dependent* |  |  |  |  |  |  |  |  |  |
| Diffused redness | 0.59 | **0.53** | **[0.29, 0.71]** | 0.24 | 0.12 | [-0.27, 0.48] | 0.36 | 0.23 | [-0.16, 0.56] |
| Color uniformity | 0.55 | **0.47** | **[0.21, 0.66]** | 0.80 | **0.71** | **[0.35, 0.86]** | 0.54 | **0.42** | **[0.06, 0.69]** |
| Dark circles, vascular | 0.18 | 0.20 | [-0.09, 0.46] | 0.46 | **0.52** | **[0.17, 0.75]** | 0.41 | **0.40** | **[0.03, 0.67]** |

We observed strong agreement between different study settings for fine texture and macro-geometric features: “Wrinkles, forehead”, “Nasolabial fold”, and “Ptosis” (Spearman’s *ρ* in range 0.68–0.95). Interestingly, high agreement of the results has been shown for the “Breakouts” parameter for SSI vs HSI and CSI vs HSI environments (ICC 0.81–0.92; SSI vs CSI ICC 0.57). At the same time, features related to the periorbital zone (“Eye bags” and “Dark circles” features) have shown low agreement when comparing the SSI and HSI study settings, indicating that this feature is highly dependent on lighting conditions and shadows, which varied between these two settings.

Comparing the two conditions where images were taken with smartphone cameras (CSI and HSI study setting), agreement remained in the range of moderate to strong for the parameters “Pigmented spots, density” (ICC = 0.88), “Nasolabial fold” (ICC = 0.85), and “Breakouts” (ICC = 0.81).

Meanwhile, AI-provided scores for fine texture and color-dependent features were significantly influenced by the smartphone-based settings compared to the high-resolution SSI settings, macro-geometric features demonstrated high stability across all devices (Table S1.2). AI-provided scores were consistent within the smartphone-based setting (CSI and HSI) regardless of the study setting.

### AI-based System vs Expert Grades

The validity of the artificial intelligence algorithms for facial skin analysis was assessed by comparing results of the AI analysis with the averaged scores assigned by the expert assessors, which served as the ground truth. To evaluate agreement between the system and experts Spearman’s rank correlation coefficient was used (Figure 1c). Chance-corrected and discretized agreement metrics, quadratic-weighted Cohen’s *κ* (Figure 1a) and the proportion of AI scores falling within one grade of the expert consensus (*±*1-grade accuracy, Figure 1b), are reported as corroborative measures of absolute agreement.

**Figure 1.**
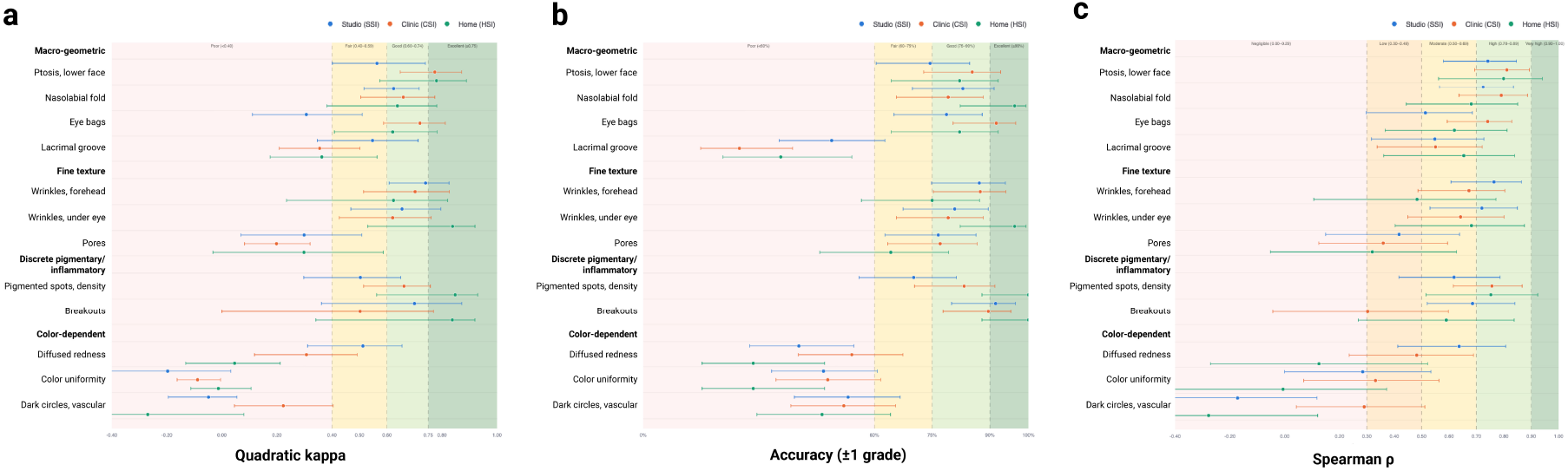
Agreement between the AI-based system and expert consensus grading across three imaging settings. (a) Quadratic-weighted Cohen’s *κ*; (b) proportion of the system’s grades falling within one grade of the expert consensus; (c) Spearman’s *ρ*. SSI, Standard Studio Imaging; CSI, Clinic Smartphone Imaging; HSI, Home Smartphone Imaging. Data presented as per-feature point estimates (dots) with whiskers indicating 95% confidence intervals.

Agreement was strongest and most setting-robust for macro-geometric features. Ptosis of the lower face maintained high rank correlation from the SSI through to HSI (*ρ* = 0.74, 0.81 and 0.80 for SSI, CSI and HSI respectively), with quadratic-weighted *κ* in the good-to-excellent range at CSI and HSI (0.77 and 0.78) and *±*1-grade accuracy of 74–85%. Nasolabial fold showed high correlation in both controlled settings (*ρ* = 0.72 and 0.79) with a moderate decline at home (*ρ* = 0.68), while *κ* remained in the “good” range (0.62–0.66) and *±*1-grade accuracy reached 96% at HSI. Eye bags showed a different pattern: correlation was only moderate in the studio setting (*ρ* = 0.51, *κ* = 0.31) but improved on smartphone acquisition (*ρ* = 0.74 and 0.62, *κ* = 0.72 and 0.62).

Forehead wrinkles showed high agreement in the studio setting (*ρ* = 0.76, *κ* = 0.74) with progressive decline across acquisition modalities (*ρ* = 0.67 at CSI, 0.48 at HSI). Under-eye wrinkles were more stable (*ρ* = 0.72, 0.64 and 0.68), with one of the highest *κ* of any feature at HSI (0.84) and *±*1-grade accuracy of 96%. Pore assessment showed only low agreement in every setting (*ρ* = 0.42, 0.36 and 0.32; *κ* = 0.30, 0.20 and 0.30), with the HSI estimate not reaching statistical significance (95% CI *−*0.05 to 0.63).

Pigmented spot density improved with smartphone acquisition, from moderate agreement in the studio setting (*ρ* = 0.62, *κ* = 0.50) to high agreement at CSI and HSI (*ρ* = 0.76 and 0.75; *κ* = 0.66 and 0.85), with *±*1-grade accuracy reaching 100% at HSI. Breakouts showed high correlation at SSI (*ρ* = 0.69, *κ* = 0.70) and moderate correlation at HSI (*ρ* = 0.59, *κ* = 0.84), with an unexplained reduction at CSI where the correlation did not reach significance (*ρ* = 0.30, 95% CI *−*0.04 to 0.60) despite *κ* of 0.50 and *±*1-grade accuracy of 90%.

Color-dependent features showed the weakest agreement overall. Diffused redness declined monotonically with decreasing acquisition control (*ρ* = 0.64, 0.48 and 0.13), with the home estimate non-significant (95% CI *−*0.27 to 0.52) and *κ* falling to 0.05. Color uniformity showed only low agreement in controlled settings (*ρ* = 0.28 and 0.33) and none at home (*ρ* = *−*0.01, 95% CI *−*0.41 to 0.37; *κ* = 0.03). Vascular dark circles showed no consistent relationship with expert grading in any setting, with point estimates falling either side of zero (*ρ* = *−*0.17, 0.29 and *−*0.28) and confidence intervals spanning zero in the studio and home settings. Lacrimal groove occupied an intermediate position, with moderate correlation across all settings (*ρ* = 0.55, 0.55 and 0.65) but fair to low *κ* (0.55, 0.36 and 0.36) and the lowest *±*1-grade accuracy of any feature at CSI (25%).

For color uniformity the three metrics diverged informatively: *±*1-grade accuracy reached 71–77% across settings while quadratic-weighted *κ* remained poor (0.24, 0.20 and 0.03) and exact agreement fell as low as 4%. Expert grades for this feature were concentrated within a narrow band of the 0–9 scale (median 5.17–6.00, IQR 0.75–1.67; Table S1.1, Figure S2.1), so agreement within one grade was largely achieved by prevalence alone while chance-corrected agreement remained low, the high-agreement, low-*κ* prevalence paradox (Feinstein and Cicchetti 1990). Pores showed the same pattern in controlled settings (*±*1-grade accuracy 77%, *κ* 0.30 and 0.20). All three metrics are therefore reported together, since none alone distinguishes concordance from concentration.

Overall, the automated system showed agreement with expert consensus sufficient to support macro-geometric features, discrete pigmentation and inflammatory lesions across all three study settings, whereas fine-texture features degraded with reduced acquisition control and color-dependent features showed limited or absent agreement irrespective of setting.

### Discussion

The aim of this study was to compare an AI-based skin analysis and grading system against clinical experts grading. Overall, our findings demonstrate that smartphone-based AI analysis of facial images supported by a platform for standardized image acquisition achieved a high concordance with the grades provided by trained assessors for macro-geometric features and pigmentation features in controlled and uncontrolled conditions. Interestingly, facial features associated with the gravitational aging and photoaging (“Ptosis” *ρ* = 0.80, “Nasolabial fold” *ρ* = 0.68, and “Pigmented spots” *ρ* = 0.75) showed high correlation even in the uncontrolled conditions of the HSI setting. This highlights that the geometric and contrast patterns underlying these facial features can be assessed with high concordance under the uncontrolled conditions typical of consumer-captured images. These findings suggest utility for hybrid and decentralized studies, provided endpoint selection is restricted to the features assessed in this work.

This study revealed systematic differences between feature classes. Macro-geometric features (ptosis and nasolabial fold) were the most concordantly graded, and notably remained so across all three settings, including participant-captured HSI images (*ρ* = 0.74–0.81 and 0.68–0.79, respectively). Eye bags, although also macro-geometric, showed greater setting dependence (*ρ* = 0.51–0.74), likely reflecting the sensitivity of periorbital contour to gaze direction and shadow geometry. Fine-texture features were intermediate and acquisition-sensitive. Color-dependent features showed the weakest agreement and deteriorated markedly under uncontrolled illumination: at HSI, color uniformity and diffused redness fell to negligible or negative rank correlation (*ρ* = *−*0.01 and 0.13), with quadratic-weighted *κ* near zero, indicating agreement no better than chance. This suggests that these features are among the most subjective or difficult to assess consistently for both expert human assessors and the AI system. For example, accurate assessment of vascular dark circles and facial redness requires cross-polarized lighting or multispectral analysis (Benvenuto-Andrade et al. 2007; Chhabra et al. 2022) to eliminate superficial tissue influence. Consequently, the analysis of these features using standard smartphone cameras without polarizing lenses is susceptible to even slight variations of lighting conditions, which is a current limitation of the remote dermatological analysis.

Grading by human experts using clinical grading scales remains one of the primary reference standards for clinical and industry applications. The results of our study reveal the inter-rater agreement among the three assessors was good to excellent across settings (median ICC 0.84–0.90), meaning that the averaged expert grade can be used as a reference standard, although agreement declined for color-dependent features under HSI conditions (color uniformity ICC = 0.40, diffused redness ICC = 0.64). Separately, the AI-based system showed high test-retest repeatability across independently captured images within each setting (median ICC 0.97–0.98). These two coefficients characterize different properties (inter-rater agreement between independent human experts, versus output stability of the system under image re-acquisition) and are therefore reported separately rather than compared directly. It is important to note that high repeatability does not mean validity as the system may generate highly reproducible scores that nonetheless would not be in concordance with expert assessment, as observed for color uniformity at HSI (test-retest ICC = 0.99 but *ρ* = *−*0.01 against expert consensus). Whether the system’s repeatability translates into sensitivity to treatment-induced longitudinal change over time was not assessed in this cross-sectional design and requires dedicated evaluation.

When comparing the smartphone images to the high-quality standardized medical-grade images, we observed a modality gap characterized by a smoothing effect. Forehead wrinkles showed a moderate to strong correlation in the two operator-captured settings (SSI and CSI study settings, *ρ* = 0.67–0.76), while the correlation for the HSI setting with unstandardized conditions decreased (*ρ* = 0.48). A similar but less prominent pattern has been shown for wrinkles under the eye. This may be the result of reduced image detail or “smoothing” in ambient and lower lighting. Medical-grade photos tend to provide more detail within the image. This highlights an important detail in remote skin analysis: macro-geometric features remain accessible, whereas fine and early aging changes such as thin superficial lines may be systematically underestimated in uncontrolled conditions relative to laboratory-grade imaging. Pore assessment, by contrast, showed only modest agreement in every setting (*ρ* = 0.32–0.42) and therefore appears limited by factors beyond acquisition standardization alone.

The performance of the AI-based system can be contextualized within the growing body of evidence in scientific literature on AI-driven facial skin analysis. Jiang et al. (2019) reported that their system achieved agreement with assessments by 12 trained experts and dermatologists, while demonstrating better reproducibility than human experts for some facial features. Subsequent validation by Flament et al. (2019) on 157 Caucasian women confirmed significant correlations between algorithm grades and dermatologist assessments for texture-related aging features. Importantly, both systems reported in previous studies (Flament et al. 2021; Flament et al. 2019; Jiang et al. 2019) and the assessed system in this study made the same fundamental observation: macro-geometric features such as sagging and nasolabial folds are the most robustly assessed from smartphone images by AI, whereas vascular and color-dependent features remain the most challenging. The correlations for ptosis (*ρ* = 0.74–0.81), nasolabial fold (*ρ* = 0.68–0.79), and pigmented spot density (*ρ* = 0.62–0.76) fall within a comparable range. However, a critical distinction of the present study is the systematic three-modality study design (medical-grade SSI, technician-captured smartphone CSI, and participant-captured smartphone HSI) applied to the same cohort and evaluated against an independent expert panel.

These findings have implications for decentralized and hybrid clinical studies, indicating that a defined subset of facial endpoints (macro-geometric aging features, discrete pigmentation, and inflammatory lesions) can be assessed from participant-captured smartphone images with agreement comparable to that obtained under standardized conditions. The high within-setting repeatability of the automated analysis (median ICC 0.97–0.98) further supports its use where consistent, operator-independent scoring is required. Conversely, endpoints dependent on absolute color fidelity require standardized illumination and should not be assigned to remote assessment without additional hardware, color-calibration measures, or expert assessment.

Limitations of the study include limited ability to accurately analyze color and light-dependent facial features, such as redness and vascular dark circles without specialized equipment using bare camera lenses. Moreover, comparing discrete grades (e.g., 0–9) with the continuous scores provided by the AI system (0–100) requires careful interpretation. However, the rank-based statistical analysis of correlation implemented in this work mitigates the scale mismatch of the results discussed above. It is worth mentioning that the studied population was narrow in terms of the skin tone. Expanding the cohort is a logical and important next step for the complex evaluation of the algorithm.

In conclusion, this study evaluated the AI-based facial analysis system against expert panel grading across three imaging modalities spanning standardized laboratory-grade acquisition to participant-captured smartphone imaging in uncontrolled environments. The system showed concordance with expert consensus sufficient to support specific endpoints (ptosis, nasolabial fold, pigmented spot density, and breakouts) under uncontrolled acquisition conditions, while endpoints dependent on absolute color fidelity require standardized illumination. Automated analysis of participant-captured facial images is therefore suitable for use as a grading tool in decentralized cosmetic and dermatological studies for a defined subset of dermatological endpoints.

## Materials and Methods

### Study Design

To assess the performance of AI-powered algorithms for facial features analysis, we evaluated the algorithms’ scores in three different experimental conditions (Figure 2): high-resolution images with standardized positioning of the face and illumination (SSI setting), smartphone images captured in a clinical setting by a trained technician (CSI setting), and smartphone images captured by the participant at home (HSI setting). A board of expert assessors evaluated the photos in each experimental setting. The results of the experts’ assessment were used as the reference standard for evaluation of the algorithms.

**Figure 2.**
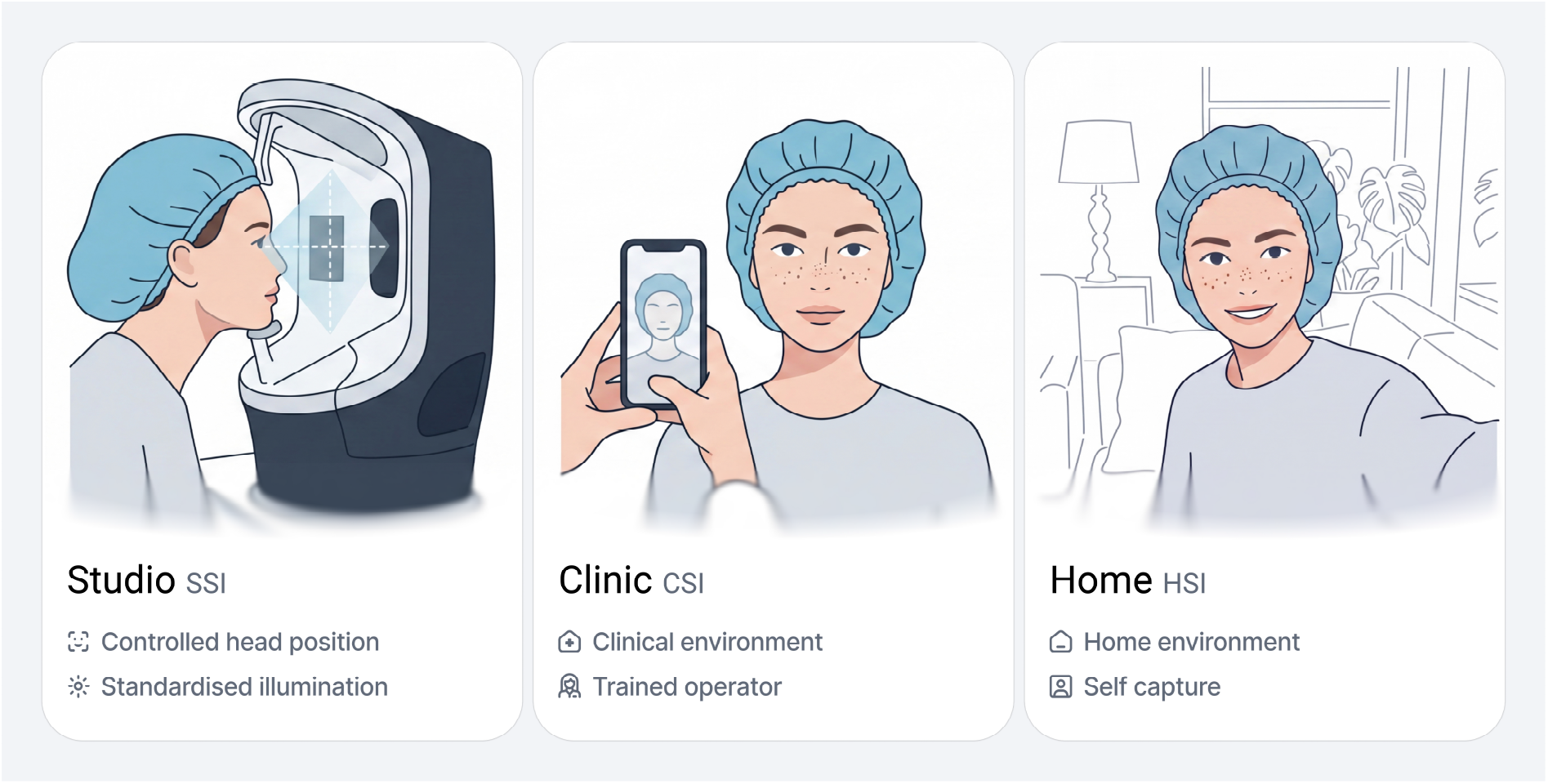
Three study settings varying in the degree of environmental control used to evaluate AI-based facial skin analysis algorithms against a panel of dermatology experts: SSI (a fully standardized clinical image-capture system), CSI (smartphone image capture by a trained technician), and HSI (smartphone image capture by the participants themselves in uncontrolled environments).

The study design was reviewed and approved by the Reading Independent Ethics Committee (protocol N°250203PE). All participants provided written informed consent for the use of their images and data in research in accordance with the Declaration of Helsinki.

### Study Participants

A total of 48 participants were enrolled in this study. The cohort was balanced in gender, comprising 26 women and 22 men (Table 5). Participant ages were broadly distributed across five decades.

**Table 5.** Demographic characteristics of study participants.

| Characteristic | No. (%) |
| --- | --- |
| Participants | 48 (100.0%) |
| *Age (years)* |  |
| 18–24 | 8 (16.7%) |
| 25–34 | 9 (18.8%) |
| 35–44 | 7 (14.6%) |
| 45–54 | 8 (16.7%) |
| 55–64 | 14 (29.2%) |
| 65+ | 2 (4.2%) |
| *Gender* |  |
| Female | 26 (54.2%) |
| Male | 22 (45.8%) |
| *Self-reported ethnicity* |  |
| Caucasian | 48 (100.0%) |
| *Fitzpatrick skin type* |  |
| II | 4 (8.3%) |
| III | 36 (75.0%) |
| IV | 8 (16.7%) |
| *Face skin type* |  |
| Oily | 5 (10.4%) |
| Mixed oily | 15 (31.3%) |
| Normal | 14 (29.2%) |
| Mixed dry | 5 (10.4%) |
| Dry | 8 (16.7%) |
| Very dry | 1 (2.1%) |

Skin phototype data indicated a predominantly homogeneous group (II–IV classes by Fitzpatrick, Table 5). The vast majority of participants were classified as Fitzpatrick phototype III (75.0%, *n* = 35), with smaller groups of phototype II (*n* = 4, 8.3%) and phototype IV (*n* = 8, 16.7%). Participants presented a diverse range of self-reported skin types. The most prevalent skin types were mixed oily (*n* = 15, 31.3%) and normal (*n* = 14, 29.2%).

Participants were eligible for inclusion in the study if they presented with at least one of the following skin concerns: wrinkles, under eye puffiness, sub-ocular fine lines, sagging, pigmented spots, dilated pores, dark circles, skin imperfections, or diffused redness. In addition, participants were required to possess a smartphone compatible with the image collection system for effective performance of the skin analysis software. Exclusion criteria included traces of facial skin irritation, pregnancy, breastfeeding, or unwillingness to take necessary precautions to avoid a pregnancy during the study.

On the day of inclusion, subjects were instructed to use only a neutral facial cleanser before the study visit. Male participants were required to be clean-shaven, with their last shave performed the evening before their inclusion in the study.

### Image Capturing

The study was conducted at the IEC clinical center (Lyon, France) under standardized environmental conditions where specific temperature and relative humidity were controlled and identical for each participant. Image capture was performed after a minimum 15-minute acclimatization period in a climate-controlled room equipped with humidity regulators. Room temperature was maintained at 20–23 °C and relative humidity at 50% *±* 10%.

#### Clinical System

High-resolution standardized facial images with standard illumination and face positioning were captured using the ColorFace^®^ (QIMA Newtone) system. The device was equipped to capture images at 24 Mpx. Participants were captured in an illustrative mode (reduced flash intensity and increased ISO) with open eyes and in a standard 60° mode to be in the same conditions as those of a smartphone, with closed eyes – one frontal photo.

One SSI image was excluded from analysis owing to an image acquisition failure, leaving 47 images available for analysis in the SSI setting.

#### Image Collection by AI-based System

To collect smartphone photos for the CSI and HSI settings, proprietary software was used for image standardization and collection. To ensure consistent image quality for reliable analysis of the images, Live Image Quality Assurance (LIQA^®^) was used. LIQA^®^ enables standardized positioning and visibility of the face in the image, as well as more standardized illumination. An internal tool for smartphone image collection was employed, compatible with the following smartphone models: Apple iPhone 8 and newer, Samsung Galaxy S20 and newer, and Xiaomi Mi 9, Note 9 and newer. Smartphone camera resolution varied substantially across device models (median 48 Mpx, range 12–200 Mpx), reflecting the heterogeneous consumer device pool typical of HSI data collection. The resolution of the images collected was at least 4032 *×* 3024 pixels (12 Mpx).

For the CSI setting image collection, each of the participants was photographed wearing a hair cap to ensure full face visibility. The image was captured by a trained technician in the standardized clinical environment on the day of the clinical assessment. The frontal photo of each participant was taken in three repetitions.

For the HSI setting, participants were instructed to take a set of photos (frontal, left side, and right side) in three repetitions upon returning home, wearing a hair cap to ensure full visibility of the face.

Overall, for each of the study settings (capture device and environment), we collected 48 images for the SSI setting in one repetition, 48 images for the CSI setting in three repetitions, and 28 images for the HSI setting in three repetitions.

### Clinical Scoring

To compare the performance of our algorithms to the severity of skin features, we utilized expert clinical grading as the reference standard. A panel of three trained assessors was assembled. Each expert annotated a frontal photo of each participant for each study setting (SSI, CSI, and HSI). Experts annotated these images in the digital grading system (IEC Scoring system) on the calibrated screen in a randomized order.

A total of 13 facial features were graded by trained assessors. Grading was performed with the help of standardized grading scales (Table 6). Skin Aging Atlases (Bazin and Flament 2010) were used to assess forehead wrinkles, eye bags, under eye wrinkles, nasolabial fold, ptosis, and density of pigmentary spots. QIMA Newtone scales were used to assess pores severity, vascular dark circles and pigmented dark circles. The IEC scoring system in 10 grades (0 to 9) was used to assess lacrimal grooves, diffused redness of the skin, and homogeneity of skin pigmentation (color uniformity). A scale by Haut.AI was used for the assessment of breakouts.

**Table 6.** Photographic grading scales and ranges for female and male participants used to evaluate the AI-based skin analysis system across 13 facial features.

| Facial feature | Scale source | Definition | Range, male | Range, female |
| --- | --- | --- | --- | --- |
| *Macro-geometric* |  |  |  |  |
| Ptoxis, lower face | Skin Ageing Atlas | Sagging severity of the lower part of the chin | 0–7 | 0–5 |
| Nasolabial fold | Skin Ageing Atlas | Depth of the fold between the base of the nose and lips | 0–6 | 0–5 |
| Eye bags | Skin Ageing Atlas | Bag of the lower eyelid | 0–5 | 0–7 |
| Lacrimal groove | IEC scale | Depth of the groove in the area under the internal eye corner along the nose | 0–9 | 0–9 |
| *Fine-texture* |  |  |  |  |
| Wrinkles, forehead | Skin Ageing Atlas | Depth of transverse wrinkles on forehead | 0–6 | 0–5 |
| Wrinkles, under eye | Skin Ageing Atlas | Depth of folds at malar area below crow’s feet | 0–6 | 0–5 |
| Pores | Qima-Newton scale | Size of pores in the area close to the nose wing | 0–5 | 0–5 |
| *Discrete pigmentary / inflammatory* |  |  |  |  |
| Pigmented spots, density | Skin Ageing Atlas | Number of spots per area unit on the cheek | 0–5 | 0–7 |
| Breakouts | Haut.AI scale | Presence and amount of pimples and inflammation | 0–5 | 0–5 |
| *Color-dependent* |  |  |  |  |
| Diffused redness | IEC scale | Red color intensity at the cheek region | 0–9 | 0–9 |
| Color uniformity | IEC scale | Skin color homogeneity on the whole face | 0–9 | 0–9 |
| Dark circles, vascular | Qima-Newton scale | Vascular dark circles visibility under the eye | 0–7 | 0–7 |
| Dark circles, pigmentary | Qima-Newton scale | Pigmented dark circles visibility under the eye | 0–9 | 0–9 |

### AI-based System

Facial features were assessed with the use of a proprietary AI system (Haut.AI, Tallinn, Estonia). A set of computer vision algorithms corresponding to the clinically assessed facial features was applied to grade the facial images.

At its core, the AI-based facial analysis system relies on deep convolutional neural networks (CNNs) which have been trained on a dataset containing more than 3 million data points. The training dataset is highly diversified, presenting a full range of skin tones, genders, and ages, ensuring fair performance across the different groups and mitigating bias. The system’s supervised learning paradigm allowed manual annotations from domain knowledge experts with biomedical and dermatological backgrounds.

In order to ensure image standardization, a Live Image Quality Assurance (LIQA^®^) system was integrated as a part of the user experience. This system directs users in real time to ensure optimal standardization of illumination and head posture for analysis. Pre-processing of images also included anonymization with the help of Skin Atlas^®^(Georgievskaya et al. 2024a) by masking non-skin pixels in the image. This approach helps protect users’ sensitive personal information.

The system provides a Face Metrics score in a continuous range from 0 to 100, where 100 represents absence of any flaws (perfect condition without concerns) and 0 is the most severe manifestation of the parameter.

### Data Analysis and Visualization

In order to assess a concordance of the clinical grading by human experts (referred to as reliability), the intraclass correlation coefficient was used as a two-way mixed model for average measures (consistency type), or ICC(3, *k*), following the model-, type-, and definition-based selection framework of Koo and Li (2016). Test-retest repeatability, which quantifies the stability of the AI-based system’s output across three independently captured photographs of the same participant within a setting and is referred to in the text as repeatability, was calculated using ICC(3, *k*) as a two-way mixed model for average measures (consistency type). At the same time, ICC(3, 1) was used as a two-way mixed model for a single measure (absolute agreement) to assess reproducibility for both experts’ annotation and the AI-based system for skin analysis between different settings.

In order to explore the relationships between various variables, we calculated non-parametric correlation coefficients: Spearman’s *ρ* accompanied by 95% confidence intervals (CI). Data are presented as the median and interquartile range (IQR), if not indicated otherwise. To detect performance differences across multiple related groups, a Friedman’s test was used, followed by a post-hoc Dunn’s test with Bonferroni correction for multiple comparisons. Spearman’s *ρ* was pre-specified as the primary agreement metric, since relative severity ordering, coefficients were interpreted using the benchmarks of Mukaka (2012). Quadratic-weighted Cohen’s *κ* was calculated on the ordinal scales and interpreted after Cicchetti (1994). *±*1-grade accuracy was calculated as the proportion of participants whose automated grade fell within one grade of the expert consensus. It was adopted as the threshold from Lorenc et al. (2021); the colored zones in the corresponding figures are descriptive rather than validated thresholds to assess performance against. We considered the differences statistically significant at the level of *p ≤* 0.05. All calculations were performed using a custom Python application based on NumPy and pandas (Python 3.13).

Because the automated score is inverted relative to the clinical scales (0 = most severe for expert grading versus 0 = most severe for the automated output), automated scores were reoriented prior to analysis so that higher values highlight greater severity in both systems. Positive coefficients therefore indicate concordance between the automated system and expert consensus, and negative coefficients indicate a relationship in the opposite direction.

## Declarations

### Author contributions

Conceptualization: AG, DD, GS; Data curation: DD, TT, LB; Formal analysis: DD, TT; Funding acquisition: AG, KK; Investigation: LB, EC; Methodology: DD, GS, TT, AG; Project administration: DD, LB; Resources: AG, KK, EC; Software: AG, KK, TT; Supervision: AG, GS; Validation: DD, TT; Visualization: DD; Writing – original draft: DD; Writing – review & editing: DD, TT, KK, LB, EC, GS, AG.

### Ethics approval and consent to participate

The study was approved by the Reading Independent Ethics Committee (Woodley, Reading, United Kingdom), study reference IEC 250203. The study was conducted in accordance with the ethical principles of the Declaration of Helsinki. Written informed consent was obtained from all participants prior to enrolment.

### Data availability

The facial images acquired in this study constitute personal data under the EU General Data Protection Regulation and cannot be shared. The de-identified datasets underlying the reported analyses (expert grades from each assessor, automated scores for each image, and the derived agreement statistics) are available from the corresponding author on reasonable request.

### Use of generative AI

Not applicable

## References

Bazin R, Flament F. Skin aging atlas. Volume 2, Asian type. Paris: Med’Com; 2010.

Benvenuto-Andrade C, Dusza SW, Agero ALC, Scope A, Rajadhyaksha M, Halpern AC, et al. Differences between polarized light dermoscopy and immersion contact dermoscopy for the evaluation of skin lesions. Arch Dermatol. 2007;143(3):329–38.

Bhor U, Pande S. Scoring systems in dermatology. Indian J Dermatol Venereol Leprol. 2006;72:315.

Chekanov K, Danko D, Tlyachev T, Kiselev K, Hagens R, Georgievskaya A. State-of-the-art in skain fluorescent photography for cosmetic and skincare research: from molecular spectra to AI image analysis. Life. 2024;14(10):1271. https://www.mdpi.com/2075-1729/14/10/1271

Chhabra N, Khare S, Sachdev D. Dermoscopy as an efficient aid to diagnose pigmentary and vascular component of periorbital melanosis: a cross-sectional study. J Cosmet Dermatol. 2022;21(11):5880–6.

Choy SP, Kim BJ, Paolino A, Tan WR, Lim SML, Seo J, et al. Systematic review of deep learning image analyses for the diagnosis and monitoring of skin disease. NPJ Digit Med. 2023;6(1):180.

Cicchetti DV. Guidelines, criteria, and rules of thumb for evaluating normed and standardized assessment instruments in psychology. Psychol Assess. 1994;6(4):284–90.

Dehon H, Brédart S. An ‘other-race’ effect in age estimation from faces. Perception. 2001;30(9):1107–13.

Escalé-Besa A, Vidal-Alaball J, Catalina QM, Gracia VHG, Marin-Gomez FX, Fuster-Casanovas A. The use of artificial intelligence for skin disease diagnosis in primary care settings: a systematic review. Healthcare. 2024;12(12):1192. https://www.mdpi.com/2227-9032/12/12/1192

Feinstein AR, Cicchetti DV. High agreement but low kappa: I. The problems of two paradoxes. J Clin Epidemiol. 1990;43(6):543–9.

Flament F, Hofmann M, Roo E, Raimbault-Gerard C, Chuberre B, Zhang Y, et al. An automatic procedure that grades some facial skin structural signs: agreements and validation with clinical assessments made by dermatologists. Int J Cosmet Sci. 2019;41(5):472–8.

Flament F, Lee YW, Lee DH, Passeron T, Zhang Y, Jiang R, et al. The continuous development of a complete and objective automatic grading system of facial signs from selfie pictures: Asian validation study and application to women of three ethnic origins, differently aged. Skin Res Technol. 2021;27(2):183–90.

Gaurav V, Agrawal S, Najeeb A, Ahuja R, Saurabh S, Gupta S. Advancements in dermatological imaging modalities. Indian Dermatol Online J. 2024;15(2):278.

George PA, Hole GJ. Factors influencing the accuracy of age estimates of unfamiliar faces. Perception. 1995;24(9):1059– 73.

Georgievskaya A, Tlyachev T, Danko D, Chekanov K, Corstjens H. How artificial intelligence adopts human biases: the case of cosmetic skincare industry. AI Ethics. 2025;5(1):105–15.

Georgievskaya A, Tlyachev T, Golodyayev A, Kiryanov R, Kiselev K. Method and system for anonymizing facial images. US Patent US12039768B2; 2024a. https://patents.google.com/patent/US12039768B2/en

Georgievskaya A, Tlyachev T, Kiselev K, Hillebrand G, Chekanov K, Danko D, et al. Predicting human chronological age via AI analysis of dorsal hand versus facial images: a study in a cohort of Indian females. Exp Dermatol. 2024b;33(3):e15045.

Ilic N, Prescott A, Erolin C, Peter M. Representation in medical illustration: the impact of model bias in a dermatology pilot study. J Vis Commun Med. 2022;45(4):253–62.

Jiang R, Kezele I, Levinshtein A, Flament F, Zhang J, Elmoznino E, et al. A new procedure, free from human assessment that automatically grades some facial skin structural signs. Comparison with assessments by experts, using referential atlases of skin ageing. Int J Cosmet Sci. 2019;41(1):67–78.

Koo TK, Li MY. A guideline of selecting and reporting intraclass correlation coefficients for reliability research. J Chiropr Med. 2016;15(2):155–63.

Lee AKW, Chan LKW, Lee CH, Bohórquez JMC, Haykal D, Wan J, et al. Artificial intelligence application in diagnosing, classifying, localizing, detecting and estimation the severity of skin condition in aesthetic medicine: a review. Dermatol Rev. 2025;6(1):e70015.

Lei C, Dang K, Song S, Wang Z, Chew SP, Bian R, et al. AI-assisted facial analysis in healthcare: from disease detection to comprehensive management. Patterns. 2025;6(2). https://www.cell.com/patterns/abstract/S2666-3899(25)00023-6

Lorenc ZP, Jones D, Kim J, Gwak HM, Batham S, Vachon G. Validating a series of photonumeric rating scales for use in facial aesthetics using statistical analysis of intra- and inter-rater reliability. Aesthet Surg J Open Forum. 2021;3(4):ojab039.

Mukaka MM. Statistics corner: a guide to appropriate use of correlation coefficient in medical research. Malawi Med J. 2012;24(3):69–71.

Neuse WHG, Neumann NJ, Lehmann P, Jansen T, Plewig G. The history of photography in dermatology: milestones from the roots to the 20th century. Arch Dermatol. 1996;132(12):1492–8.

Skea ZC, Newlands R, Gillies K. Exploring non-retention in clinical trials: a meta-ethnographic synthesis of studies reporting participant reasons for drop out. BMJ Open. 2019;9(6):e021959. https://bmjopen.bmj.com/content/9/6/e021959

Tobar MdPB, Clemann S, Hagens R, Pagel-Wolff S, Hoppe S, Behm P, et al. Skinly: a novel handheld IoT device for validating biophysical skin characteristics. Skin Res Technol. 2024;30(3):e13613.

Wilczok D. Deep learning and generative artificial intelligence in aging research and healthy longevity medicine. Aging. 2025;17(1):251–75.

