## Supplementary materials S1, S2 for "Comparison of Facial Feature Grading by an AI-Based System Versus Human Expert Grading on Images Under Standardized Clinical and At-Home Settings"

---

**Contents**

S1 Distribution of expert grades and automated scores across study settings (Tables S1.1–S1.2)
S2 The system's vs experts' grades (Figure S2.1)

---

### S1 – Distribution of Expert Grades and Automated Scores Across Study Settings

**Table S1.1.** Comparison of median grades from the expert panel for 13 facial features across three study settings (SSI – standardized environment, high-resolution images; CSI – technician-captured smartphone images; HSI – participant-captured smartphone images). Data are presented as median and interquartile range (IQR). Statistical significance was assessed using the Friedman test ($\chi^2$) for overall differences across the three groups, followed by Dunn's post-hoc test with Bonferroni correction for pairwise comparisons ($z$-score). Bold values indicate statistical significance. Dashes: post-hoc testing not performed.

| | SSI | | CSI | | HSI | | Friedman | | SSI vs CSI | | SSI vs HSI | | CSI vs HSI | |
|---|---|---|---|---|---|---|---|---|---|---|---|---|---|---|
| Facial feature | Median | IQR | Median | IQR | Median | IQR | $\chi^2(2)$ | $p$ | $z$ | $p$ | $z$ | $p$ | $z$ | $p$ |
| *Macro-geometric* | | | | | | | | | | | | | | |
| Ptosis, lower face | 0.67 | 2.08 | 1.17 | 2.75 | 0.83 | 2.17 | **17.94** | **0.0001** | −1.19 | 0.7014 | −0.32 | 1.0000 | −0.72 | 1.0000 |
| Nasolabial fold | 1.67 | 2.42 | 1.00 | 2.08 | 1.50 | 1.42 | **7.23** | **0.0270** | 0.83 | 1.0000 | −0.94 | 1.0000 | −0.22 | 1.0000 |
| Eye bags | 1.00 | 1.42 | 1.50 | 2.00 | 0.67 | 1.75 | **8.97** | **0.0113** | −1.68 | 0.2796 | 0.54 | 1.0000 | 1.99 | 0.1415 |
| Lacrimal groove | 2.83 | 4.00 | 3.00 | 2.67 | 2.33 | 2.08 | 2.29 | 0.3176 | – | – | – | – | – | – |
| *Fine-texture* | | | | | | | | | | | | | | |
| Wrinkles, forehead | 2.00 | 2.42 | 1.50 | 2.00 | 0.67 | 1.83 | 5.86 | 0.0534 | – | – | – | – | – | – |
| Wrinkles, under eye | 1.00 | 2.00 | 1.00 | 1.92 | 0.33 | 1.08 | 3.98 | 0.1371 | – | – | – | – | – | – |
| Pores | 1.67 | 1.42 | 1.33 | 1.08 | 1.50 | 2.33 | 1.22 | 0.5425 | – | – | – | – | – | – |
| *Discrete pigmentary / inflammatory* | | | | | | | | | | | | | | |
| Pigmented spots, density | 1.33 | 2.00 | 1.00 | 1.00 | 0.67 | 1.08 | **10.02** | **0.0067** | 0.04 | 0.9696 | **2.65** | **0.0080** | 1.68 | 0.0935 |
| Breakouts | 0.33 | 1.33 | 0.33 | 1.00 | 0.33 | 0.67 | **6.68** | **0.0355** | 0.52 | 1.0000 | 0.84 | 1.0000 | 0.40 | 1.0000 |
| *Color-dependent* | | | | | | | | | | | | | | |
| Diffused redness | 3.17 | 3.25 | 2.17 | 2.33 | 1.83 | 2.50 | 2.63 | 0.2680 | – | – | – | – | – | – |
| Color uniformity | 5.17 | 1.67 | 5.67 | 1.33 | 6.00 | 0.75 | **7.01** | **0.0301** | **−2.33** | **0.0200** | **−2.70** | **0.0069** | 0.00 | 1.0000 |
| Dark circles, vascular | 2.00 | 2.00 | 3.00 | 1.75 | 2.00 | 2.42 | **28.00** | **0.0000** | −1.78 | 0.2243 | 0.26 | 1.0000 | 1.79 | 0.2190 |
| Dark circles, pigmentary | 1.33 | 4.50 | 0.33 | 3.67 | 0.33 | 0.58 | **20.05** | **0.0000** | 1.68 | 0.0925 | **2.36** | **0.0182** | 0.00 | 1.0000 |

#### Detailed Analysis of the Experts' Grading

To assess whether the image acquisition method influenced clinical assessment, expert consensus grades were analyzed using the non-parametric Friedman test. As detailed in Table S1.1, no statistically significant differences were observed between the CSI (technician-captured) and HSI (participant-captured) settings for any of the measured facial features (Dunn's post-hoc, $p > 0.05$ for all significant parameters). This confirms that participant-captured images are non-inferior to technician-captured images for the purpose of expert grading.

However, acquisition modality (SSI high-resolution imaging vs smartphone CSI and HSI) influenced grading for specific color-dependent and discrete pigmentary and inflammatory features. Significant differences were observed between the SSI baseline and the HSI setting for "Pigmented spots, density" ($p$ = 0.008), "Dark circles, pigmented" ($p$ = 0.0182), and "Color uniformity" ($p$ = 0.0069). In these cases, the high-resolution, controlled lighting of the SSI system likely allowed experts to visualize subtle pigmentation variations that were less apparent in smartphone images, leading to divergent grading distributions. Conversely, "Lacrimal groove" and fine texture features showed no significant differences in grading distributions across any setting ($p > 0.05$), supporting the robustness of analysis across different camera types.

**Table S1.2.** Comparison of median scores generated by the AI-based system across three study settings for facial features (SSI – high-resolution images captured in a standardized setting; CSI – smartphone images captured by a trained technician in a standardized setting; HSI – smartphone images captured by participants in their homes). Data are presented as median and interquartile range (IQR). Statistical significance was assessed using the Friedman test ($\chi^2$) for overall differences across the three groups, followed by Dunn's post-hoc test with Bonferroni correction for pairwise comparisons ($z$-score). Bold values indicate statistical significance. Dashes: post-hoc testing not performed.

| | SSI | | CSI | | HSI | | Friedman | | SSI vs CSI | | SSI vs HSI | | CSI vs HSI | |
|---|---|---|---|---|---|---|---|---|---|---|---|---|---|---|
| Facial feature | Median | IQR | Median | IQR | Median | IQR | $\chi^2(2)$ | $p$ | $z$ | $p$ | $z$ | $p$ | $z$ | $p$ |
| *Macro-geometric* | | | | | | | | | | | | | | |
| Ptosis, lower face | 79.0 | 43.5 | 69.0 | 34.5 | 90.7 | 46.8 | 3.06 | 0.2163 | – | – | – | – | – | – |
| Nasolabial fold | 2.0 | 2.0 | 2.0 | 1.7 | 1.8 | 1.0 | **9.91** | **0.0070** | 0.00 | 0.1059 | 0.01 | 0.9952 | **2.41** | **0.0158** |
| Eye bags | 93.0 | 15.5 | 89.0 | 26.9 | 87.2 | 22.8 | 3.39 | 0.1840 | – | – | – | – | – | – |
| Lacrimal groove | 66.0 | 27.5 | 55.5 | 30.1 | 52.7 | 19.6 | **12.07** | **0.0024** | 1.81 | 0.0698 | **2.01** | **0.0449** | 0.00 | 1.0000 |
| *Fine-texture* | | | | | | | | | | | | | | |
| Wrinkles, forehead | 65.0 | 35.0 | 77.2 | 23.3 | 88.0 | 18.8 | **7.60** | **0.0223** | −0.19 | 0.8460 | **−2.57** | **0.0100** | −1.51 | 0.1310 |
| Wrinkles, under eye | 62.0 | 36.5 | 61.7 | 20.8 | 81.0 | 13.8 | 2.89 | 0.2359 | – | – | – | – | – | – |
| Pores | 83.8 | 10.4 | 79.0 | 21.5 | 83.3 | 20.3 | 0.73 | 0.6946 | – | – | – | – | – | – |
| *Discrete pigmentary / inflammatory* | | | | | | | | | | | | | | |
| Pigmented spots, density | 86.7 | 18.3 | 83.0 | 12.0 | 88.2 | 11.6 | 5.85 | 0.0536 | – | – | – | – | – | – |
| Breakouts | 79.0 | 18.0 | 93.5 | 8.0 | 89.0 | 12.6 | **15.45** | **0.0004** | **−4.27** | **0.0000** | −1.28 | 0.2010 | 1.55 | 0.1214 |
| *Color-dependent* | | | | | | | | | | | | | | |
| Diffused redness | 54.7 | 20.3 | 49.0 | 22.5 | 53.0 | 14.2 | 1.81 | 0.4039 | – | – | – | – | – | – |
| Color uniformity | 55.0 | 16.0 | 57.7 | 11.3 | 57.7 | 7.0 | 1.73 | 0.4204 | – | – | – | – | – | – |
| Dark circles, vascular | 66.7 | 14.9 | 72.0 | 17.5 | 70.2 | 7.6 | **19.57** | **0.0001** | **3.54** | **0.0004** | 1.18 | 0.2378 | −0.89 | 0.3735 |

#### Detailed Analysis of the System's Scores

To evaluate the impact of imaging setup (including resolution and lighting) on automated scoring, the distributions of scores were analyzed using the Friedman test. As shown in Table S1.2, the acquisition modality significantly influenced the absolute scoring of fine-texture features, indicative of a "smoothing effect" inherent to smartphone photography.

Significant differences were observed for fine texture features. For "Wrinkles, forehead" the median score was significantly lower in the high-resolution SSI setting (median = 65.0) compared to the HSI setting (median = 88.0, $p = 0.0100$). Similarly, "Breakouts" were scored as more severe in the SSI setting (median = 79.0) compared to the smartphone-based CSI setting (median = 93.5, $p < 0.0001$). This suggests that the higher resolution and contrast of the clinical system allow for the detection of subtle defects that may be missed by the lower resolution of smartphone sensors. Conversely, macro-geometric features showed high distributional stability across devices, with no statistically significant differences for "Ptosis" or "Eye bags". No significant difference was observed for "Diffused redness" either, although the absence of a distributional shift for this feature should not be read as evidence of valid grading, given its poor agreement with expert consensus (main text, Figure 1).

When comparing the two smartphone-based settings (CSI vs HSI), the system demonstrated high distributional stability. No significant differences were found for 11 of the 12 facial features, with only "Nasolabial folds" showing a statistically significant but numerically minor distributional shift ($p = 0.0158$). This confirms that while absolute scores may differ from medical-grade imaging, the AI metrics remain consistent within the smartphone domain regardless of the operator.

### S2 – The System's vs Experts' Grades

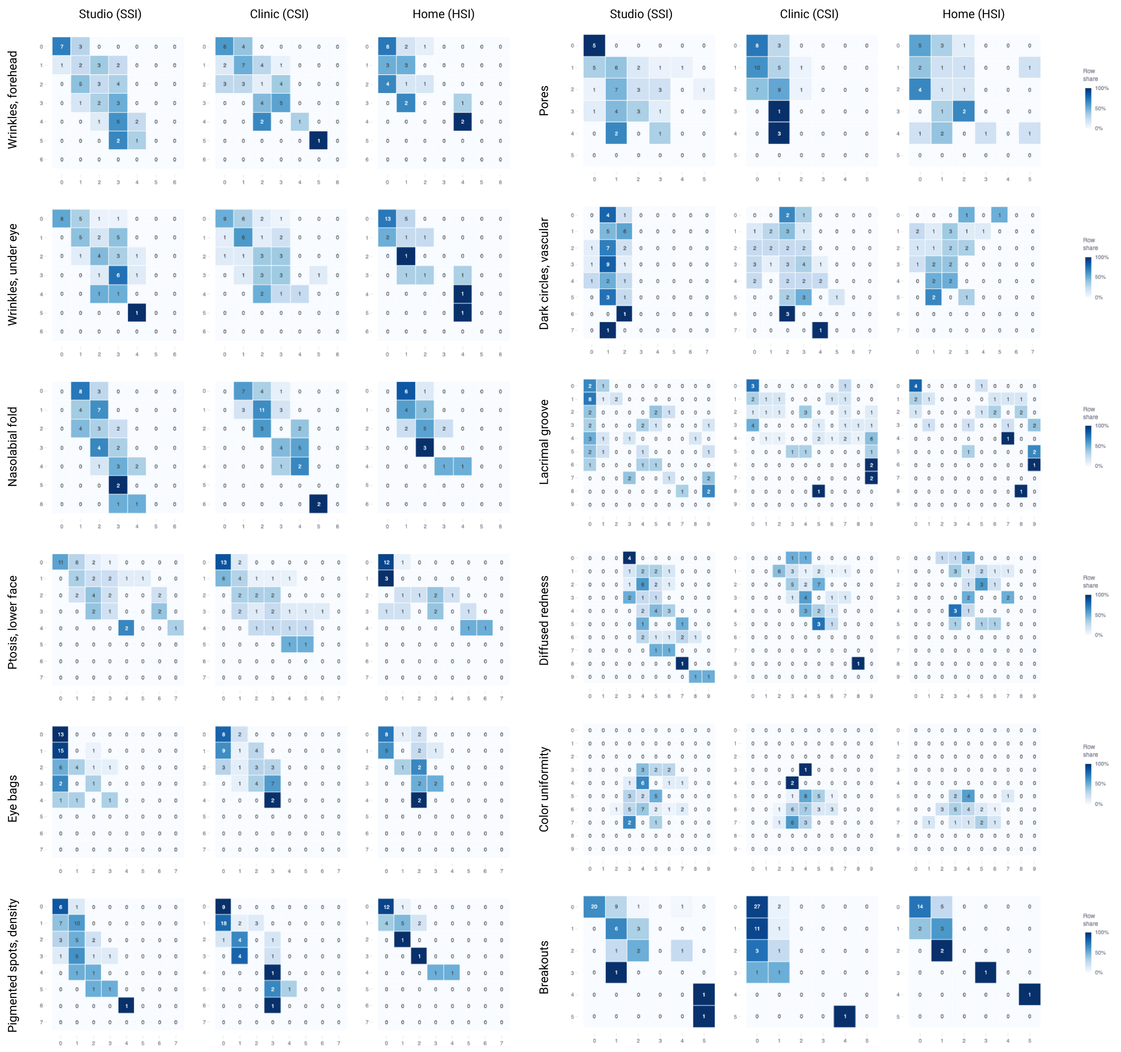


**Figure S2.1.** Confusion matrices comparing grades assigned by the AI-based system with the averaged expert consensus grade for 12 facial features across three study settings (SSI – high-resolution images captured in a highly standardized environment; CSI – smartphone images captured by a trained technician in a standardized environment; HSI – smartphone images captured by participants in their homes). Rows indicate the expert consensus grade and columns the grade derived from the automated score after mapping onto the applicable expert scale; cell values are participant counts and color intensity denotes the proportion of each row (row share).

To characterize the structure underlying the summary agreement metrics, the joint distribution of expert consensus grades and automated grades was examined for each facial feature and study setting (Figure S2.1). Whereas rank correlation, chance-corrected agreement and $\pm 1$-grade accuracy each reduce this distribution to a single value, the matrices show how disagreements were distributed relative to the diagonal and how much of the available scale range each feature occupied.

For macro-geometric and for discrete pigmentary and inflammatory features, observations were concentrated on or immediately adjacent to the diagonal in all settings, indicating that disagreements were predominantly of one grade rather than gross misclassification. Exact agreement was highest for breakouts (62%, 58% and 64% for SSI, CSI and HSI respectively) and for pigmented spot density at HSI

(64%).

Where disagreements occurred, the off-diagonal mass lay predominantly below the diagonal, indicating that automated grades tended to fall below the expert consensus (most apparent for eye bags at SSI, where expert grades of 1 to 3 were repeatedly returned as grade 0). Such an offset reduces chance-corrected agreement while leaving rank correlation comparatively intact, consistent with the metrics for that feature ($\rho$ = 0.51, $\kappa$ = 0.31).

The matrices also clarify the divergence between $\pm$1-grade accuracy and chance-corrected agreement described in the main text. For color uniformity, expert grades occupied only the central portion of the 0–9 scale (IQR 0.75 grades at HSI, Table S1.1), so adjacent-grade agreement was readily achieved while exact agreement remained low (21%, 31% and 4%). Vascular dark circles and lacrimal groove instead showed observations dispersed across grade categories with limited diagonal alignment; lacrimal groove had the lowest exact agreement of any feature (6%, 10% and 21%) despite moderate rank correlation, indicating that ordering was preserved more successfully than absolute grade.
